# Developing and validating the Nepalese Abuse Assessment Screen (N-AAS) for identifying domestic violence among pregnant women in Nepal

**DOI:** 10.1101/2023.09.25.23296107

**Authors:** Pratibha Manandhar, Pratibha Chalise, Poonam Rishal, Jacquelyn Campbell, Lena Henriksen, Jennifer J. Infanti, Sunil Kumar Joshi, Mirjam Lukasse, Kunta Devi Pun, Berit Schei, Katarina Swahnberg, the ADVANCE 2 study team

## Abstract

This study culturally adapted and validated a Nepalese version of the Abuse Assessment Screen (AAS) tool for identifying domestic violence among pregnant women in Nepal, creating the Nepalese Abuse Assessment Screen (N-AAS). International and national topic experts reviewed the initial N-AAS version using the Delphi method, and pregnant women participated in cognitive interviews, providing feedback on the N-AAS as user experts. Subsequent pre-testing of a comprehensive questionnaire, which included the translated version of the N-AAS, occurred in two tertiary care hospitals using an electronic format known as Color-Coded Audio Computer-Assisted Self-Interview (C-ACASI). We assessed the content validity index, compared the concurrent validity of the N-AAS with the gold standard interview, estimated the prevalence of domestic violence from two hospitals, and calculated the Kappa coefficient. The reliability of the entire questionnaire was also evaluated through a test-retest analysis, with content validity rated as “good to excellent” by topic and user experts and high test-retest reliability (91.2–98.9%), indicating consistency across questionnaires completed at two different time points, with 12% of participants reporting any form of violence. The N-AAS demonstrated ≥91.7% specificity for all forms of abuse, accurately identifying non-abuse cases. In addition, moderate to excellent sensitivity was observed for emotional abuse (52.5%) and physical abuse since marriage (50%), while sensitivity for physical abuse in the past 12 months was 100%. Thus, the N-AAS demonstrated reliable test-retest results with a good Kappa coefficient and specificity, as well as showing excellent sensitivity for detecting recent physical abuse and moderate sensitivity for detecting emotional abuse and physical abuse since marriage. Because cultural context often leads women to normalize and tolerate abuse from spouses and family members and women are thus reluctant to report abuse, the results imply that the N-AAS can serve as a valuable screening tool for domestic abuse in antenatal care settings in Nepal.

## Introduction

Domestic violence (DV) is a global concern that encompasses various forms of harm inflicted within familial relationships. The Nepal Ministry of Law and Justice has defined DV as “any physical, mental, sexual, or economic harm perpetrated by one person on another with whom he or she has a family relationship, including acts of reprimand or emotional harm”[1]. DV during pregnancy, including intimate partner violence (IPV), is a serious concern that can result in maternal death and morbidity, either directly or indirectly [2], and it is associated with adverse maternal health and pregnancy outcomes, such as placental abruption, miscarriage, ante-partum hemorrhage, and inadequate weight gain, as well as poor perinatal outcomes, such as low birth weight and preterm delivery [3–5].

Prevalence studies of DV and IPV against pregnant women from around the world show a wide range of reported figures. According to a multi-country study on women’s health and domestic violence against women conducted by the World Health Organization (WHO), physical violence during pregnancy by a male intimate partner affects between 1% and 28% of women globally [6]. Variability is even more pronounced when considering developing versus industrialized countries, with prevalence rates of violence of female partners during pregnancy ranging from 3.8–31.7% in developing countries compared 3.4–11% in industrialized countries [7]. A more recent review of 80 individual studies from different countries revealed prevalence rates of IPV during pregnancy ranging from 1.5–66.9% in low- and middle-income countries (LMICs) [8]. Moreover, psychological violence emerged as the most prevalent form of violence during the perinatal period, with rates ranging from 1% in Sweden to 81% in South Africa, while physical violence prevalence ranged from 0.4% in Sweden to 60.6% in Uganda [8]. In the two hospital locations where we conducted the current study, prior research by our study team demonstrated that 21% of pregnant women attending routine antenatal care (ANC) reported experiencing all forms of DV [9]. Another study among pregnant women attending ANC in a hospital in eastern Nepal found that 3.5% of women reported physical DV during their current pregnancy [10].

The significant variability in reported IPV and DV prevalence can be attributed to several factors, including differences in measurement instruments, varying definitions of violence, and diverse cultural contexts. Additionally, the sensitive nature of this topic can impact how participants respond to surveys. Therefore, it is imperative to develop and validate culturally and contextually relevant measurement instruments to provide a more accurate understanding of the complex dynamics surrounding violence during pregnancy.

Preventing and reducing the harms of DV requires the involvement of the health sector, multi-sectoral organizations, awareness campaigns, and economic empowerment, with ANC services being particularly important because most women attend them during their reproductive lives [10]. In 2016, 84% of women who gave birth in the previous five years in Nepal reported attending at least one ANC visit during their pregnancies, an increase of 25% from 2011, and 69% of Nepalese women attended four or more ANC visits throughout their pregnancies [11].

Several studies have introduced tools, instruments, or questionnaires for detecting pregnancy-related DV in Nepal [10, 12, 13]. The Abuse Assessment Screen (AAS), originally developed in the United States [14], has been validated in different countries, including Turkey, Greece, and Spain, demonstrating its effectiveness as a simple tool [15–17]. However, formal validation of the AAS is still lacking in LMICs, including Nepal. Therefore, the objective of this study was to develop, to adapt culturally, and to validate a linguistically relevant version of the AAS specifically for routine clinical use in Nepal. This version is called the Nepalese Abuse Assessment Screen (N-AAS), whose test-retest reliability we also endeavored to analyze.

## Materials and methods

This validation study was carried out over one year, from December 2021 to November 2022, with the main steps outlined in Fig 1.

**Fig 1.**
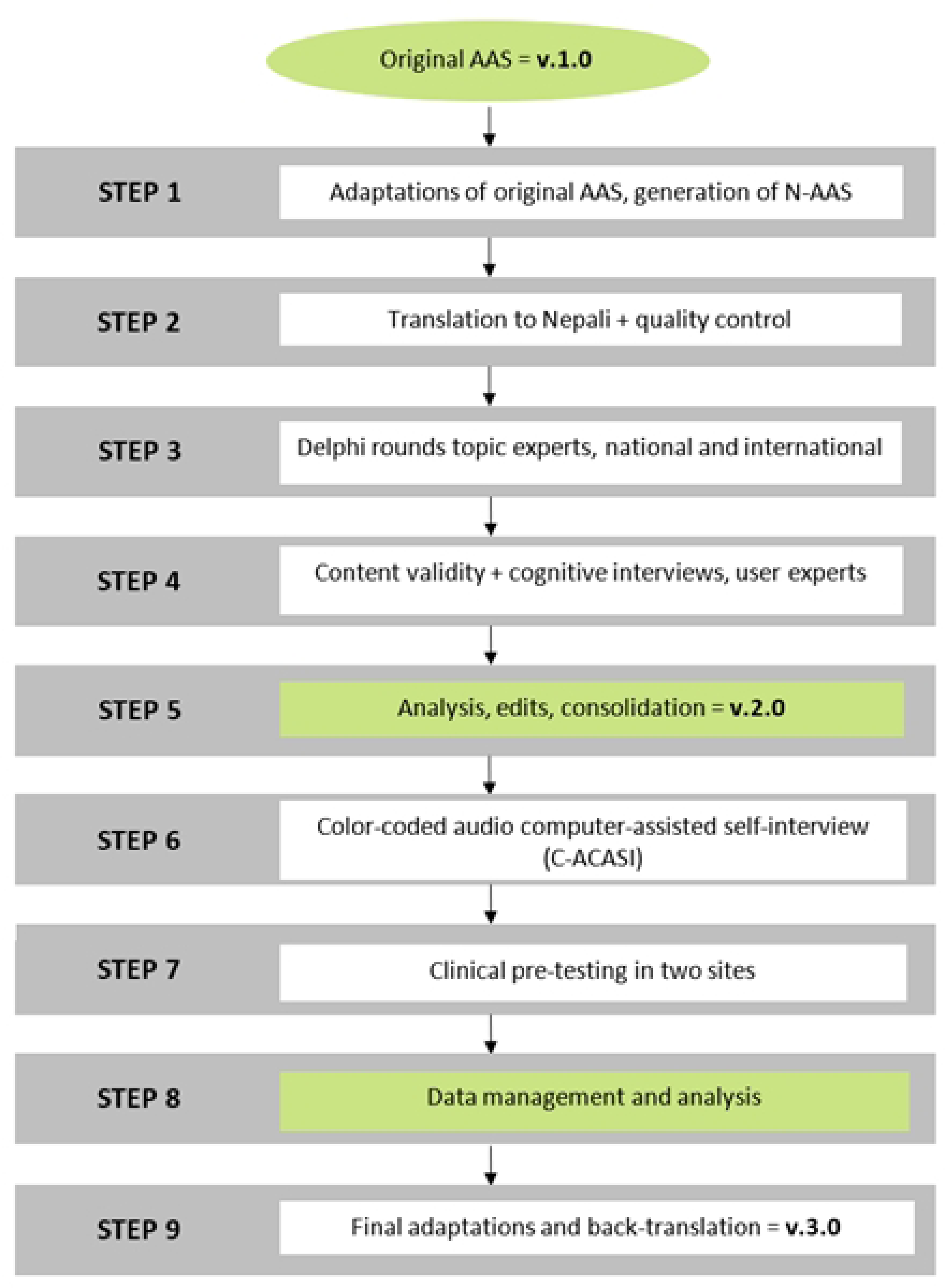
Step-by-step process of developing and validating the Nepalese Abuse Assessment Screen (N-AAS).

### Step 1: Adapting the original AAS and generating the N-AAS

The AAS is an effective and short tool for detecting DV during pregnancy in clinical practice, and it has demonstrated effectiveness in identifying DV among pregnant women in developed countries [18] and in developing countries [19–21]. In our study team’s previous prevalence and qualitative studies, conducted at the same sites as the present study, the AAS was used as a data collection tool [10, 22]. However, we did not include the “body mapping” component in these studies due to cultural sensitivity and social taboos. In addition, the AAS question about “fear in the family” was identified as inconsistent with the Nepalese context, where “fear” can be interpreted as a sign of respect for family members. Therefore, these two components were excluded from the initial version of the N-AAS.

Furthermore, the qualitative study [22] revealed that demanding physical labor and food denial were perceived as forms of physical and emotional mistreatment by pregnant women’s family members. Based on these findings, two new questions were added to the original AAS, specifically targeting these pregnancy- and culturally specific types of emotional violence [10, 22], resulting in the N-AAS.

A few additional adjustments were made to enhance the clarity and focus of the initial version of the N-AAS: we revised the first question to concentrate on DV and IPV by asking about experiences since marriage or living with a partner, rather than encompassing lifetime abuse. In addition, we separated the original AAS question on emotional and physical abuse into distinct questions to assess better these specific forms of abuse. We also expanded the answer choices to include mother-in-law, father-in-law, and other family members, while excluding the option for stranger. More detailed information regarding these modifications can be found in Table 1.

**Table 1.** Comparison between the original Abuse Assessment Screen (AAS) and the Modified Nepalese Version (N-AAS), Validated in the Study.

| Original Abuse Assessment Screen (AAS) |  | Nepalese Abuse Assessment Screen (N-AAS) |  |
| --- | --- | --- | --- |
| Question | Answer option(s) | Question | Answer option(s) |
| 1. Have you ever been emotionally or physically abused by your partner or someone important to you? | Yes/No<br>If YES, who? (Circle all that apply):<br>Husband<br>Ex-husband<br>Boyfriend<br>Stranger<br>Other<br>Multiple<br>Total # of times: | 1. Since you've been married or living with a partner, have you been emotionally abused by someone in the family?<br>*(Examples: Emotional abuse includes such acts as saying or doing something to humiliate you intentionally in front of others, the use of abusive words directed at you, threatening to hurt or harm you and your child/children, insulting you, deliberately making you feel bad about yourself, prohibiting you from visiting your mother's home, etc.)* | Yes/No<br>If YES, by whom? (Choose one or more options):<br>Husband<br>Ex-husband<br>Boyfriend<br>Mother-in-law<br>Father-in-law<br>Other family member<br>How many times?<br>Once or twice<br>Several times (3–5 times)<br>Many times (more than 5 times) |
|  |  | 2. Since you've been married or living with a partner, have you been physically abused by someone in the family?<br>*(Examples: slapped, burned on purpose, kicked, dragged, or otherwise intentionally hurt, had your hair pulled, spat on, struck with objects, or otherwise intentionally hurt)* | Yes/No<br>If YES, by whom? (Choose one or more options):<br>Husband<br>Ex-husband<br>Boyfriend<br>Mother-in-law<br>Father-in-law<br>Other family member<br>How many times?<br>Once or twice<br>Several times (3–5 times)<br>Many times (more than 5 times) |
| 2. Within the last year, have you ever been hit, slapped, kicked, or otherwise physically hurt by someone? | Yes/No<br>If YES, who? (Circle all that apply):<br>Husband<br>Ex-husband<br>Boyfriend | 3. Within the last year, have you been hit, slapped, kicked, had your hair pulled, spat on, struck with an object, or otherwise physically hurt by someone in the family? | Yes/No<br>If YES, by whom? (Choose one or more options):<br>Husband<br>Ex-husband<br>Boyfriend<br>Mother-in-law |
|  | Stranger<br>Other<br>Multiple<br>Total # of times: |  | Father-in-law<br>Other family member<br>How many times?<br>Once or twice<br>Several times (3–5 times)<br>Many times (more than 5 times) |
| 3. Since you've been pregnant, have you been slapped, kicked, or otherwise physically hurt by someone? | Yes/No<br>If YES, who? (Circle all that apply):<br>Husband<br>Ex-husband<br>Boyfriend<br>Stranger<br>Other<br>Multiple<br>*Supplementary body mapping exercise:*<br>*The person being assessed is provided with a blank body outline or a diagram of the human body. They are asked to mark the area of injury on the body map.* | 4. Since you've been pregnant, have you been hit, slapped, kicked, had your hair pulled, spat on, struck with an object, or otherwise physically hurt by someone in the family? | Yes/No<br>If YES, by whom? (Choose one or more options):<br>Husband<br>Ex-husband<br>Boyfriend<br>Mother-in-law<br>Father-in-law<br>Other family member<br>How many times?<br>Once or twice<br>Several times (3–5 times)<br>Many times (more than 5 times) |
|  |  | 5. Since you've been pregnant, have you been forced by someone in the family to do heavy physical work despite feeling unwell or exhausted?<br>*(Examples: forced heavy physical work includes activities such as being made to carry a 20 liter of water jar or load of hay, forced to sit for a long period of time while washing clothes, lift sacks of rice, prolonged walking, etc.)* | Yes/No<br>If YES, by whom? (Choose one or more options):<br>Husband<br>Ex-husband<br>Boyfriend<br>Mother-in-law<br>Father-in-law<br>Other family member<br>How many times?<br>Once or twice<br>Several times (3–5 times)<br>Many times (more than 5 times) |
|  |  | 6. Since you've been pregnant, has someone in the family given you insufficient or no food to eat despite the availability of food and money in the household?<br>*(Examples: not providing nutritious food such as eggs, fish, meat, fruits or hiding those foods)* | Yes/No<br>If YES, by whom? (Choose one or more options):<br>Husband<br>Ex-husband<br>Boyfriend<br>Mother-in-law |
|  |  |  | Father-in-law<br>Other family member<br>How many times?<br>Once or twice<br>Several times (3–5 times)<br>Many times (more than 5 times) |
| 4. Within the last year, has anyone forced you to have sexual activities? | Yes/No<br>If YES, who? (Circle all that apply):<br>Husband<br>Ex-husband<br>Boyfriend<br>Stranger<br>Other<br>Multiple<br>Total # of times: | 7. Within the last year, has anyone in the family forced you to have sexual activities without your consent?<br>*(Examples: physically forced you to have sexual intercourse or attempting to try when you did not want to, touching or trying to touch your private parts, using sexual slurs, forcing you with threats or in any other way to perform sexual acts you did not want to, or otherwise coercing you into reproductive acts?)* | Yes/No<br>If YES, by whom? (Choose one or more options):<br>Husband<br>Ex-husband<br>Boyfriend<br>Mother-in-law<br>Father-in-law<br>Other family member<br>How many times?<br>Once or twice<br>Several times (3–5 times)<br>Many times (more than 5 times) |
| 5. Are you afraid of your partner or anyone listed above? | Yes/No |  |  |

#### Study questionnaire

In this study, the N-AAS was validated as a component (Section D) of a comprehensive questionnaire designed to assess various aspects of a safe pregnancy. The comprehensive questionnaire, developed for our study team’s subsequent research phase, a randomized controlled trial, comprised eight sections and 110 questions in total. The sections included: Section A: Recent pregnancy, Section B: Participant and family information, Section C: Food access, Section D: Abuse in family relationships (assessed with the N-AAS), Section E: Support seeking and other coping strategies, Section F: Attitudes towards family behaviors and norms, Section G: Self-reported general health status, and Section H: Safety promoting strategies.

### Step 2: Translation to Nepali and quality control

The 110-item questionnaire was translated from English into Nepali by a professional translator and psychologist fluent in both languages. Back translation of the abuse questions (only the N-AAS) was performed by a native English and Nepali speaker.

### Step 3: Delphi rounds with topic experts to evaluate content validity

To evaluate the content validity of the N-AAS, the following steps were conducted iteratively using the Delphi method [23] from December 2021 to April 2022. Initially, the study team selected a review panel consisting of nine international and nine national topic experts, including nurses, psychologists, researchers, clinicians, and members of scientific networks from various organizations. To gather their input, a N-AAS review form was prepared and sent to the experts, who were asked to assess the individual items based on their relevance, understandability, and whether the items were sensitive using a five-point Likert scale. A score of 0 indicated the item was irrelevant, non-understandable, and highly sensitive, whereas a score of 4 indicated the item was highly relevant, easily understandable and non-sensitive. The experts provided scores for each item and shared qualitative comments, which were carefully considered to refine the domains and their respective items. In addition, the content validity index (CVI), analyzed in step 5, was calculated based on their assessments.

### Step 4: Content validity and cognitive interviews with user experts

The user expert group consisted of 18 pregnant women aged 18 years and older, in their 12-20 weeks of gestation. This was a convenience sample recruited from two tertiary care hospitals in Bagmati province during the women’s ANC visits in May and June 2022.Twelve women had experienced DV, while six women did not.

The user experts also evaluated and rated the N-AAS items based on their relevance, understandability, and sensitivity. The CVI was then calculated based on their assessments, following the same process as the topic experts. Additionally, these participants took part in cognitive interviews using a think-aloud technique [24]. This approach aimed to assess whether the content of the N-AAS items accurately and meaningfully reflected the experiences of the interviewees. The primary focus of the interviews was to evaluate the women’s comprehension, judgment, recall and response to the questions based on their personal experiences. Throughout the interviews, the first and second authors posed probing questions to encourage reflections, and field notes were taken.

### Step 5: Analysis

Next, in an analysis phase, the N-AAS items were evaluated using the CVI from both topic and user experts. Item-level content validity index (I-CVI) scores were calculated. The scale-level content validity index/average (S-CVI/Ave) and scale-level content validity index/universal acceptance (S-CVI/UA) were determined, using the same approach for all experts. The content validity of the N-AAS was assessed in terms of relevance, understandability, and sensitivity [25]. The specific CVI scores obtained from this analysis are presented in the results section.

### Step 6: Color-Coded Audio Computer-Assisted Self-Interview (C-ACASI)

The 110-item questionnaire, which included the N-AAS, was converted into an electronic data collection format known as C-ACASI [26]. This study used two versions of the questionnaire. Initially, the questionnaire only offered a listening option. However, after a first performance assessment, the team decided to enhance the questionnaire by incorporating both reading and listening options.

### Step 7: Clinical pre-testing

#### Test-retest

Clinical pre-testing was conducted at the study team’s two collaborating hospitals, referred to as Hospital 1 and Hospital 2, from June to November 2022. Hospital 1 covers both urban and rural areas, while most participants from Hospital 2 are from urban areas. During the pre-testing period, a total of 7437 pregnant women attended ANC, of which 1180 were eligible participants (12-20 weeks of pregnancy, aged ≥18 years), with 768 participants from Hospital 1 and 412 participants from Hospital 2.

Pregnant women attending the Outpatient Department (OPD) of Obstetrics and Gynecology departments at the hospitals were referred to the data collection site by the nursing staff after their registration. At the data collection site, pregnant women were provided with invitations to participate in the study and received detailed explanations about it. Either a Research Assistant (RA) or the first or second author provided comprehensive explanations about the questionnaire, including the C-ACASI format and the use of electronic tablets to administer the study questionnaire. To establish a relationship with the participants, some general questions unrelated to the study were asked, such as about hobbies and favorite foods. Participants were assured of confidentiality and informed that some questions in the C-ACASI might be complex, sensitive, and of a personal nature. Participants were encouraged to ask for clarification if any questions were unclear, and the functioning of the ‘back’ and ‘next’ buttons on the C-ACASI was explained and demonstrated. Participants’ unique code numbers, mobile numbers and addresses were recorded in a logbook, which was stored securely in a locked cupboard when not in use. Basic information such as height, weight, last menstrual period date and hemoglobin levels were entered into the tablet by research staff prior to handing it over to the participant to complete the questionnaire. Upon completion, participants were thanked and informed that they would receive calls for a retest in two to eight weeks’ time. Participants were provided with booklets on safe pregnancy, nutrition, family planning, and non-communicable diseases.

When participants returned for the retest, they were given a brief overview of the study’s procedures again and reminded of their right to interrupt the process at any time. Upon giving consent, they were provided with a tablet to complete the questionnaire for the second time. Several reasons were given for not enrolling in the study, including medical issues, a busy OPD schedule, inability to return for follow-up, language challenges, and a lack of interest due to time constraints. A total of 488 participants were enrolled (235 from Hospital 1 and 253 from Hospital 2). The Time 1 questionnaire (test) was completed by 467 participants (221 from Hospital 1 and 246 from Hospital 2), while the remaining participants (n=21) discontinued during the C-ACASI process (see Fig 2 for recruitment and sample details).

**Fig 2.**
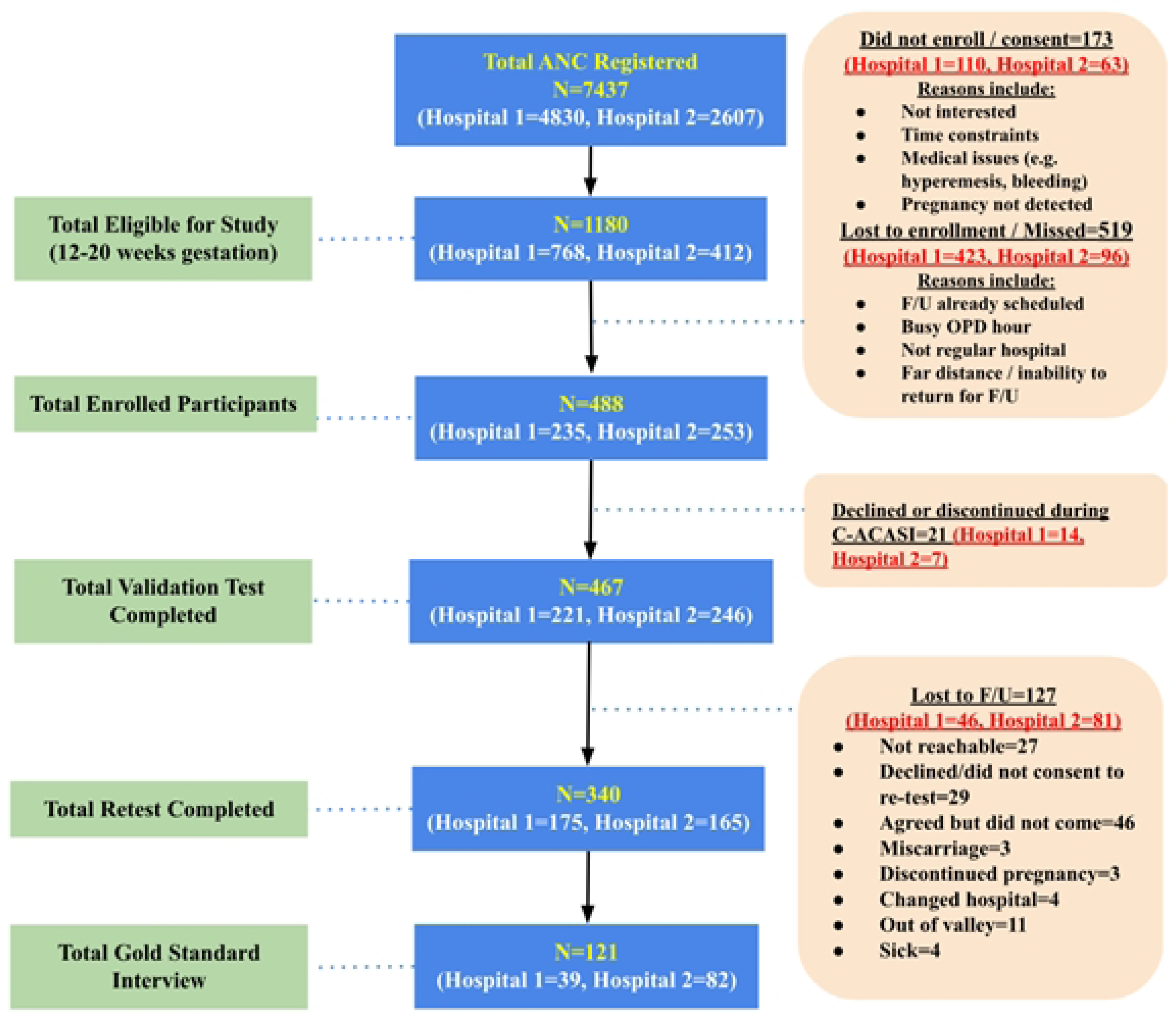
Recruitment and samples from two hospitals for clinical pre-testing (Hospital 1 and Hospital 2).

At the second time point, the retaking of the questionnaire (retest), which was held two to eight weeks after the initial test, some women chose not to participate again, and some again withdrew during the C-ACASI process. A total of 340 participants (72.8%) completed the retest. Reasons for not participating in the retest included being unreachable by mobile, agreeing but not showing up, pregnancy termination, miscarriage, switching hospitals, being out of the valley and a family member’s illness.

#### Gold standard interviews

To assess the measurement precision of the N-AAS, a total of 121 participants (39 from Hospital 1 and 82 from Hospital 2) were invited to participate in a short, structured interview known as the ‘gold standard interview’ [27]. An interview, involving face-to-face interactions, is considered the most accurate method (therefore, the ‘gold standard’) to provide results closest to the truth in this study.

Participants who answered positively to at least one of the seven N-AAS questions were invited for the interviews at a ratio of 3:1 compared to those who answered negatively. Two weeks after completing the initial questionnaire, a RA contacted the participants. The first and second authors were blinded to the participants’ DV status. Some of these women completed the interviews on a special visit for the purpose, others while attending their routine follow-up visits two to eight weeks after completing the initial questionnaire, while others did not answer the RA’s phone call but made a surprise visit to the hospitals for the interviews (S1 Fig). The interviews were conducted in a private room and audio recorded. On average, DV-negative cases took around seven minutes to complete the interview, while DV-positive cases took 20-40 minutes.

**S1 Fig.**
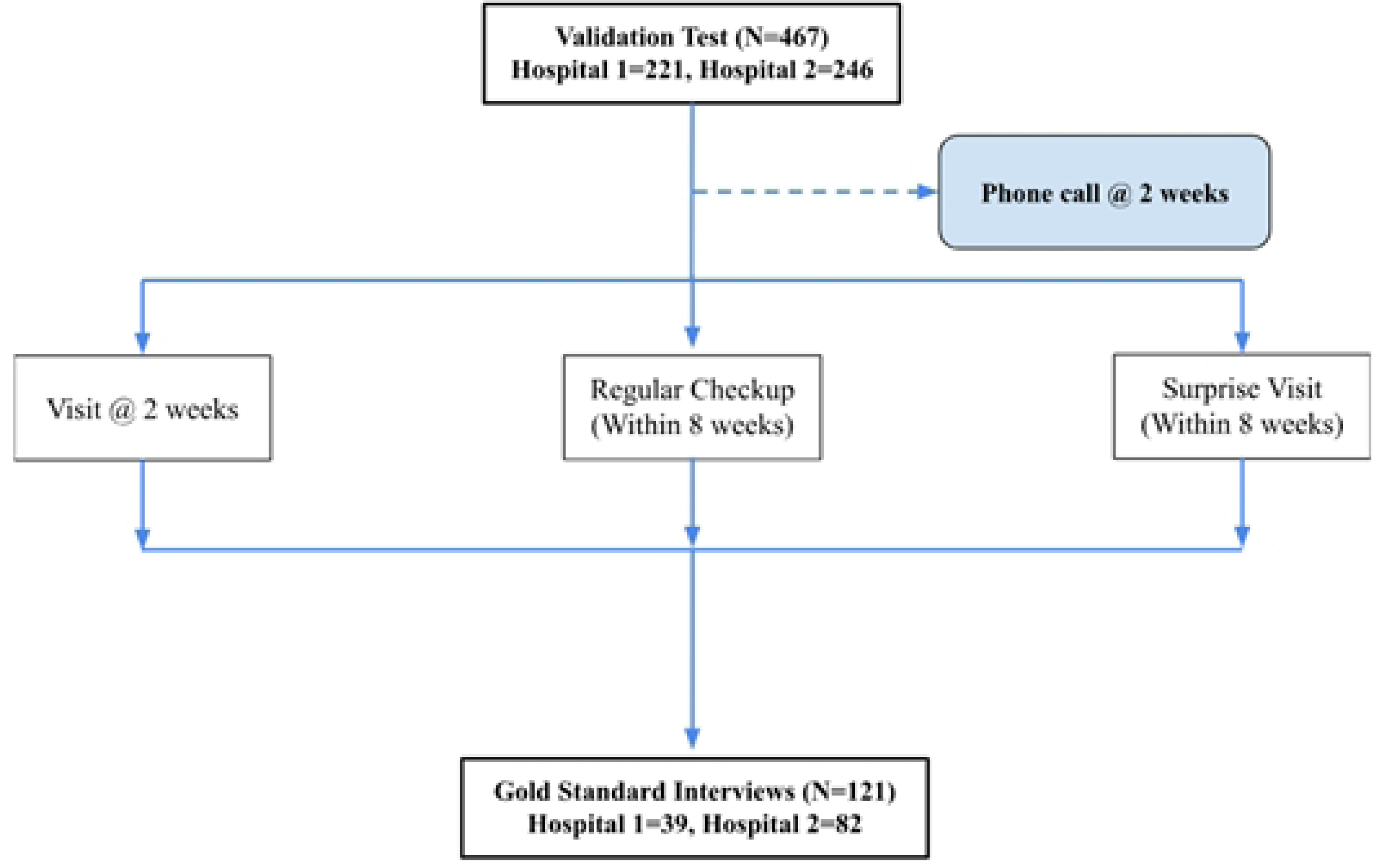
Recruitment process and samples for gold standard interviews from two hospitals (Hospital 1 and Hospital 2).

#### Ethical considerations

Prior to collecting data, formal ethical approvals were obtained from The Nepal Health Research Council, Nepal (Ref: 2395) and The Regional Committee for Medical and Health Research Ethics of Central Norway (REK) (Ref: 178092). The study comprised a two-step consent process. First, verbal agreement for participation was obtained, which involved explaining the study objective to the participants and asking if they were willing to receive further and more detailed information. The second step was obtaining electronic consent in the C-ACASI. The authors also acquired informed consent for recording the gold standard interviews. Participants were informed that their participation was voluntary and that they had the right to withdraw from the study at any point without any impact on their future ANC.

### Step 8: Data management and analysis

SPSS version 26 was used to analyze the data. The association between the interview sample and the entire sample from the two hospitals was examined using the chi-square test and Fisher’s exact in bivariate analysis, with a p-value of ≤0.05 considered statistically significant. The concurrent validity items i.e., prevalence, sensitivity, specificity, positive and negative predictive values, and likelihood ratios were assessed [28], compared with the gold standard interview [29]. Cohen’s kappa test of concordance was used to establish reliability [30]. The 110-item questionnaire was assessed in percentages for test-retest reliability after administering the same questionnaire twice over two to eight weeks.

### Step 9: Final adaptations and back translation

Upon reflecting on the results of the clinical pretesting, the study team made a few final modifications to the N-AAS items, such as expanding the introduction to the N-AAS questions and adding illustrative examples in several questions to clarify the different types and forms of violence.

The final version of the Nepali language N-AAS was back-translated to English by a fluent speaker of both languages. The study team then compared the two English versions. The team’s two PhD students (first and second authors) along with their supervisors evaluated the forward and backward translations to ascertain whether the true meaning had been captured.

## Results

### Content validity

Based on the I-CVI calculations, the scores obtained for the N-AAS were good (0.92–0.96) in terms of relevance and understandability for I-CVI, S-CVI/Ave, and S-CVI/U. In terms of sensitivity, where lowest scores are good levels, the score obtained for the N-AAS was 0.6, which is satisfactory. The range of CVI obtained good scores for relevance (0.57–0.98), understandability (0.57–1), and sensitivity (0–0.14) in the Delphi round with topic experts and among the user expert group (Table 2).

**Table 2.** Content Validity Assessment: Delphi and User Expert Group Item Scale Values.

|  | Relevance |  |  | Understandability |  |  |  | Sensitivity |  |  |  |
| --- | --- | --- | --- | --- | --- | --- | --- | --- | --- | --- | --- |
|  | IE | NE | UE | IE | NE<br>Delphi<br>1 | NE<br>Delphi<br>2 | UE | IE | NE<br>Delphi<br>1 | NE<br>Delphi<br>2 | UE |
| **I-CVI** | 0.96 | 0.94 | 0.98 | 0.92 | 0.90 | 0.96 | 1 | 0.61 | 0.77 | 0.66 | 0.14 |
| **S-CVI/Ave** | 0.96 | 0.94 | 0.98 | 0.92 | 0.90 | 0.96 | 1 | 0.66 | 0.77 | 0.60 | 0.14 |
| **S-CVI/UA** | 0.70 | 0.57 | 0.85 | 0.57 | 0.57 | 0.85 | 1 | 0 | 0 | 0 | 0 |

Notes: IE, International Experts (N=9); NE, National Experts (N=9); UE, User Experts (N=18); I-CVI, Item-Level Content Validity Index; S-CVI/Ave=Scale-Level Content Validity Index/Average; S-CVI/UA, Scale-Level Content Validity Index/Universal Agreement.

The qualitative feedback obtained from the international topic experts highlighted important considerations, such as asking questions with care and respect and assisting women who may be experiencing emotional distress, including shame, worry, anxiety, and other distressing emotions. The experts also emphasized the importance of employing trained interviewers to ensure more accurate answers. In response to recommendations from the national experts, culturally adapted examples of abuse, such as hair pulling, spitting on, and forcing to lift heavy sacks of rice and hay, were added to the N-AAS (Table 1).

The user experts, pregnant women in the study setting, emphasized the understandability, relevance, acceptability, feasibility, and cultural appropriateness of the N-AAS questions, and they noted that healthcare settings can provide a safe space for individuals who have experienced violence to disclose their experiences [31]. However, they underscored that participants in studies about DV must be assured that their responses to questionnaires will be published only in a manner that guarantees their anonymity.

### Background characteristics

Table 3 shows the background characteristics of the participants in the clinical pre-testing phase of validation, showing the differences in total participants (n=461) vs. interview participants (n=121) and in Hospital 1 (n=221) vs. Hospital 2 (n=246). The largest proportion (44%) of study participants were aged 25 to 29 years, with no difference between the two samples (p=0.72) and two hospitals (p=0.155). In addition, most (62%) participants were of Brahmin/Chhetri (upper caste) ethnic group, with a similar distribution in the total and interview samples (p=0.24). However, in Hospital 1, the largest proportion of participants were from the Janajati (Indigenous) ethnic group (43.4%), whereas in Hospital 2, most (65%) were from the Brahmin/Chhetri group (p<0.001). Participants’ civil status was equally distributed in both samples (p=0.14), but not equally distributed in both hospitals (p=0.019). Further, there was no significant difference in the distribution of the samples in both categories according to their independent income (p=0.286, p=0.399) with the majority of participants not having an independent income in either hospital, type of family (p=0.67, p= 0.577), with just over half in both hospitals living in “joint” families, and approximately half in each hospital experiencing their first pregnancy (p=0.64, 0= 0.853).

**Table 3.** Background Characteristics Comparing the Interview and Total Sample and Comparing Participating Hospitals.

|  |  | Total sample<br>N=467 | Interview sample<br>n=121 | p-value | Hospital 1<br>n=221 | Hospital 2<br>n=246 | p-value |
| --- | --- | --- | --- | --- | --- | --- | --- |
|  |  | n (%) | n (%) |  | n (%) | n (%) |  |
| **Age (years)** | < 20 | 19 (4) | 6(5) | 0.72 | 12(5.4) | 7(2.8) | 0.155 |
|  | 20–24 | 126 (27) | 28(23.1) |  | 66(29.9) | 60(24.4) |  |
|  | 25–29 | 205 (44) | 57(47.1) |  | 88(39.8) | 117(47.6) |  |
|  | 30–34 | 101 (22) | 25(20.7) |  | 50(22.6) | 51(20.7) |  |
|  | 35 or more | 15 (3) | 5(4.1) |  | 5(2.3) | 11(4.5) |  |
| **Ethnicity** | Janajati<sup>a</sup> | 159 (34) | 32(26.4) | 0.24 | 96(43.4) | 63(25.6) | <0.001 |
|  | Brahmin/Chhetri<sup>b</sup> | 252 (54) | 75(62.0) |  | 92(41.6) | 160(65) |  |
|  | Dalit<sup>c</sup> | 20 (4) | 7(5.8) |  | 15(6.8) | 5(2) |  |
|  | Madhesi<sup>d</sup> | 6 (1.2) | 1(0.8) |  | 0 | 6(2.4) |  |
|  | Muslim<sup>e</sup> | 2 (0.42) | 0 |  | 0 | 2(0.8) |  |
|  | Others<sup>f</sup> | 28 (6) | 6(5) |  | 18(8.1) | 10(4.1) |  |
| **Settlement** | Rural | 121 (26) | 22(18.2) | 0.02 | 106(48) | 15(6.1) | <0.001 |
|  | Urban | 346(74) | 99(81.8) |  | 115(52) | 231(93.9) |  |
| **Education** | Illiterate | 20(4) | 4(3.3) | 0.76 | 6(2.7) | 14(5.7) | 0.036 |
|  | Informal education | 28(6) | 8(6.6) |  | 12(5.4) | 16(6.5) |  |
|  | Upto class 8 | 59(13) | 19(15.7) |  | 29(13.1) | 30(12.2) |  |
|  | Class 9–12 | 197(41) | 50(41.3) |  | 108(48.9) | 89(36.2) |  |
|  | >12 | 163(36) | 40(33.1) |  | 66(29.9) | 97(39.4) |  |
| **Independent income** | Yes | 121 (25.9) | 46(38.1) | 0.286 | 89(40.3) | 109(44.3) | 0.399 |
|  | No | 346 (74.1) | 152(43.9) |  | 132(59.7) | 137(55.7) |  |
| **Type of family** | Nuclear | 212(45) | 57(47.1) | 0.67 | 97(43.9) | 115(46.7) | 0.577 |
|  | Joint | 255 (55) | 64(52.9) |  | 124(56.1) | 131(53.3) |  |
| **Civil status** | Married, living with husband | 431(92) | 110 (90.9) | 0.14 | 211(95.5) | 220(89.4) | 0.019 |
|  | Married, not living with husband | 36(8) | 11(9.1) |  | 10(4.6) | 26(10.3) |  |
| **Primi gravida** | Yes | 227(48.6<br>) | 61(50.4) | 0.64 | 106(48) | 121(49.2) | 0.853 |
|  | No | 240(51.4<br>) | 60(49.6) |  | 115(52) | 125(50.8) |  |

<sup>a</sup>Janajati-Indigenous groups with varying degrees of opportunity and access to social mobility, including some with little or no social mobility.
<sup>b</sup>Brahmin/Chhetri (Upper castes)-Brahmin are traditionally the most privileged groups in the social hierarchy.
<sup>c</sup>Dalit-The most oppressed social class
<sup>d</sup>Madhesi-Groups residing/originating from the Terai region.
<sup>e</sup> Muslim-Group from Muslim religion
<sup>f</sup> Others- Groups other than the abovementioned categories

### Prevalence of violence

Any form of violence was reported by 12% of participants with almost similar proportions in both study sites (12.7% and 11.4%). Among all types of violence, emotional abuse was most prevalent (10.1%) and food deprivation was least prevalent (0.6%) (Table 4).

**Table 4.** Prevalence of Abuse.

| N-AAS items |  | N=467 | Hospital 1<br>n=221 (47%) | Hospital 2<br>n=246 (53%) |
| --- | --- | --- | --- | --- |
|  |  | N (%) | n (%) | n (%) |
| **Emotional abuse** | Since you've been married or living with a partner as married, have you been emotionally abused by someone in the family? | 47(10.1) | 21(9.5) | 26(10.6) |
| **Physical abuse (since marriage)** | Since you've been married or living with a partner as married, have you been physically abused by someone in the family? | 14(3) | 6(2.7) | 8(3.3) |
| **Physical abuse (past 12 months)** | Within the last year, have you been hit, slapped, kicked, spat on, or struck with an object; had your hair pulled; or otherwise physically abused by someone in the family? | 12(2.6) | 2(0.9) | 10(4.1) |
| **Physical abuse (since pregnancy)** | Since you've been pregnant, have you been hit, slapped, kicked, spat on, or struck with an object; had your hair pulled; or otherwise physically abused by someone in the family? | 7(1.5) | 3(1.4) | 4(1.6) |
| **Forced to do heavy work** | Since you've been pregnant, have you been forced by someone in the family to perform heavy physical work despite feeling unwell or exhausted? | 9(1.9) | 4(1.8) | 5(2) |
| **Food deprivation** | Since you've been pregnant, has someone in the family given you insufficient or no food to eat despite food being available in the house? | 3(0.6) | 1(0.5) | 2(0.8) |
| **Sexual abuse (past 12 months)** | Within the last year, has anyone in the family forced you into sexual activity without your consent? | 9(1.9) | 2(0.9) | 7(2.8) |
| **Yes, to any type of abuse** |  | 56(12) | 28(12.7) | 28(11.4) |

### Concurrent validity

The N-AAS demonstrated a high range of specificity (91.7–100%) across all types of abuse in both listening-only and listening and reading versions, where the sensitivity of the former in identifying emotional abuse was 34.4% and the latter 52.5%. Regarding physical abuse, the N-AAS showed a sensitivity of 50% for abuse since marriage and 100% for abuse within the last 12 months (see Table 5 for detailed results).

**Table 5.** Concurrent Validity Analysis of the Seven N-AAS Abuse Items Compared to Interview Gold Standard.

|  | EA | PA since marriage | PA at 12m | PA since pregnant | FHW | FD | SA 12m | EA+FD+ FHW | PA+<sup>a</sup> |
| --- | --- | --- | --- | --- | --- | --- | --- | --- | --- |
| **N-AAS listening only version<sup>b</sup>, n=68 (% and likelihood ratio)** |  |  |  |  |  |  |  |  |  |
| Sensitivity | 34.4 | 33.3 | 20 | 0 | 28.6 | 33.3 | 14.3 | 37.5 | 64.3 |
| Specificity | 91.7 | 98.4 | 100 | 95.5 | 98.4 | 100 | 98.4 | 91.7 | 94.4 |
| Pre-test probability | 47.1 | 8.8 | 7.4 | 2.9 | 10.3 | 4.4 | 10.3 | 47.1 | 11.6 |
| (prevalence) |  |  |  |  |  |  |  |  |  |
| Positive predictive value | 78.6 | 66.7 | 100 | 0 | 66.7 | 100 | 50 | 80 | 60 |
| Negative predictive value | 61.1 | 93.8 | 94 | 96.9 | 92.3 | 97 | 90.9 | 62.3 | 95.3 |
| Likelihood ratio | 7.31 | 6.715 | 5.4 | 0.183 | 6.01 | 6.605 | 2.099 | 8.78 | 26.21 |
| **N-AAS listening and reading version<sup>c</sup>, n=53 (% and likelihood ratio)** |  |  |  |  |  |  |  |  |  |
| Sensitivity | 52.5 | 50 | 100 | 0 | 25 | 0 | 20 | 55 | 85.7 |
| Specificity | 84.6 | 89.4 | 97.9 | 98.1 | 97.8 | 100 | 93.6 | 84.6 | 91.3 |
| Pre-test probability (prevalence) | 75.5 | 11.3 | 9.4 | 1.9 | 15.1 | 9.4 | 9.6 | 75.7 | 13.2 |
| Positive predictive value | 91.3 | 37.5 | 83.3 | 0 | 66.7 | 0 | 25 | 91.7 | 60 |
| Negative predictive value | 36.7 | 93.3 | 100 | 98.1 | 88 | 90.6 | 91.7 | 37.9 | 97.7 |
| Likelihood ratio | 6.03 | 4.80 | 27.71 | 0.03 | 4.46 | 0 | 0.88 | 6.78 | 18.41 |

EA, Emotional abuse; PA, Physical abuse; FHW, Forced to do heavy work; FD, Food deprivation; SA, sexual abuse.
<sup>a</sup>PA+, Physical abuse since marriage + Physical abuse past 12 months (12m) + Physical abuse since pregnant.
<sup>b</sup>The first version of the C-ACASI, consisting of listening questions only.
<sup>c</sup>The second version of the C-ACASI, including both listening and reading options.

### Test-retest reliability

The test-retest reliability measure for the N-AAS ranged from 91.2–98.9%, with the highest percentage obtained for sexual violence (98.9%) and the lowest for emotional abuse (91.2%). The Kappa coefficient was calculated for the N-AAS, showing the highest agreement for sexual abuse (0.74) and the lowest for physical abuse since pregnancy (-0.00) (S1 Table).

**S1 Table. Test–Retest Reliability of N-AAS Items (with Kappa Coefficient).**

The test-retest reliability of the entire comprehensive questionnaire was also measured (S2 Table), and the test-retest reliability range was0.27–99.4% for Section A, 81.8–100% for Section B, 81%–97.2% for Section C, 91.2–98.9% for Section D, 83.3–95.8% for Section E, 87.3–97.8% for Section F, 80.7–97.2% for Section G, and 45.8–100% for Section H.

**S2 Table. Test-Retest Reliability of Comprehensive Questionnaire (with Kappa Coefficient).**

## Discussion

The current study addresses gaps in the validation of tools for screening DV among pregnant women in LMICs. Through a comprehensive validation process, the N-AAS tool was developed as a culturally adapted simple screening tool with seven relevant questions for the Nepalese context.

### Content validity

Content validity is an as essential aspect of evaluating the appropriateness and representativeness of an assessment tool for the construct of interest [32, 33]. In this study, the content validity of the N-AAS was assessed through a rigorous iterative process involving input from a team of experts who provided feedback on the relevance, understandability, and sensitivity of the N-AAS items.

Overall, regarding the cultural adaptation of the N-AAS, the CVI demonstrated good scores, indicating the experts’ agreement on the relevance, understandability, and sensitivity of the questions. These findings align with similar observations from the validation of the AAS in Turkey [15], reinforcing the validity and potential broader utility of the N-AAS in cultures other than the US.

The insights from the user experts in our study emphasize the need to prioritize the creation of user-friendly and culturally appropriate tools within healthcare contexts, as they enable the identification and support of people experiencing DV. The N-AAS questions were reported as straightforward, reasonably sensitive, and easy to answer, making them suitable for inclusion in routine antenatal care services.

The questionnaire was administered using tablet computers and a digital technology called C-ACASI [26], an approach proven effective for collecting data on sensitive topics within ANC settings with limited privacy. Participants’ privacy was maintained, and the majority remained engaged throughout the study, indicating a satisfactory level of comfort with the questionnaire. The use of electronic data collection methods, such as C-ACASI, offers several advantages in busy outpatient departments, where time is limited. It provides a confidential data collection tool that accommodates individuals with minimal or no literacy, particularly relevant in a country with a female literacy rate of around 67% [11]. Previous studies have also highlighted the effectiveness of C-ACASI in ensuring high-yield and private responses to sensitive questions [10].

The modifications made to the listening and reading versions of the questionnaire simplified and expedited the data collection process, saving time and improving participant comprehension. By incorporating user feedback and leveraging digital technology, the N-AAS demonstrated practicality and ensured participant comfort, privacy, and inclusivity. Further, the utilization of C-ACASI not only improved the data collection process but also expanded the accessibility of the questionnaire to include a wider range of participants.

### Concurrent validity

The current investigation revealed the N-AAS had high specificity, positive predictive value, and adequate to high negative predictive value, demonstrating its overall satisfactory level of measurement accuracy. Sensitivity was higher in the reading and listening version than in the listening version only, which could be attributed to added clarity for participants who could read but who may have misheard or misunderstood while listening only. In addition, completing the questionnaire may be less burdensome for participants in the hearing and listening version because it took less time to complete, which may explain the difference in results shown in Table 5.

Another study conducted in Nepal [34] highlighted that cultural factors often lead women to accept violence from their husbands. To encourage disclosure and improve relevance and understanding, we incorporated culturally relevant examples of physical abuse and being forced to do heavy work into the N-AAS. However, disclosures in response to these questions were limited, with few instances of physical abuse and food deprivation reported, and the low disclosure rates observed in the study could potentially be attributed to the success of recent public health initiatives. For instance, increased ANC visits have allowed family members, alongside pregnant women, to receive health information from qualified specialists, and similar initiatives emphasize the importance of pregnant women consuming nutritious meals and of raising awareness about their vulnerability during pregnancy [35].

In the C-ACASI listening version, we observed lower sensitivity than the gold standard interview, where some women initially denied experiencing DV in the C-ACASI questionnaire but disclosed instances of DV during the interviews. This inconsistency suggests a potential weakness in the C-ACASI format for DV inquiry and may signal a lack awareness, understanding, or recognition of abuse among participants. This observation aligns with a qualitative study [32] on South Asian women’s DV experiences, indicating that societal pressures often discourage them from disclosing or revealing DV due to fear of family embarrassment. It is possible that some participants are unfamiliar with the term “domestic violence,” which may explain the discrepancy in responses observed in our study. Furthermore, such factors as low confidence, distrust of ethnic or social outsiders, loyalty to family, and a desire to protect family privacy and honor could have contributed to the reluctance to disclose DV. However, our study revealed false negative cases with low sensitivity in the C-ACASI listening version, underscoring the need for awareness-raising programs and interventions to combat DV, fostering greater consciousness and reducing general societal acceptance of DV.

Studies validating DV screening tools and instruments are an important contribution to addressing the complex issue, and they are crucial to developing innovative interventions that involve collaboration among legal, judicial, law enforcement, social services, and medical systems, followed by comprehensive evaluations [36]. In addition, qualitative research plays a role in gaining valuable insights into potential strategies for increasing screening rates and developing successful intervention programs, particularly from the perspective of the women themselves [37]. As such, our study, by validating a DV screening instrument, contributes to these ongoing efforts.

### Prevalence

The reported prevalence rates of DV among pregnant women in this study appeared low, with 12% for any type of abuse, compared to our prior study which documented a prevalence of 21% [10]. This disparity can primarily be attributed to the limitations imposed by our sample size in the current study. To address this limitation and obtain a more comprehensive understanding of DV prevalence, especially in terms of specific characteristics such as the type of perpetrator, we are currently conducting a larger scale randomized controlled trial (RCT) at the same study sites.

Additionally, another significant factor that may explain the low reported prevalence of DV in this study is the persistence of a patriarchal system in Nepal [22], which may discourage the reporting of DV and normalize violence perpetrated by intimate partners [38].

### Test-retest reliability

The test-retest reliability of the N-AAS was found to be high, indicating that the questions produced consistent results when administered at two different time points. The Kappa coefficient measures agreement between observations, the findings of which highlight the simplicity and effectiveness of the N-AAS as a tool for assessing DV prevalence among pregnant women in Nepal.

In a validation study of the AAS conducted in Turkey [15], the reliability coefficient was reported as KR20=0.801, whereas the Greek version of the scale yielded a reliability coefficient of 0.80 [39]. Therefore, the comparability of the N-AAS result with other studies supports it as a robust and consistent tool for measuring DV among pregnant women.

### Contributions to knowledge

This study contributes to the research methodology and evidence-base regarding validated tools for addressing DV in LMIC contexts. One significant contribution is the enhanced understanding of the cultural meanings and nuances of violence, where the inclusion of culturally specific forms of violence in the tool, such as food deprivation despite the availability of food in the house, represents a step towards improving the recognition and understanding of DV in a cultural context. Although the positive response rate to this question was low, it underscores the importance of further testing these questions in our larger clinical study.

Identifying DV in pregnancy is just one aspect of broader efforts aimed at combating all types of violence against women. Additional measures are needed to shift societal norms, raise awareness, provide education, and train healthcare providers to inquire about DV experiences. Various health promotion programs should be implemented from the national level to break the silence surrounding the disclosure of DV experiences. Continued and further preventive work in this field is important to ensure the safety and well-being of individual women who have experienced DV.

### Strengths and limitations

This study represents a pioneering validation of a clinical screening tool for DV in Nepal, the development of which and similar screening tools is crucial for obtaining better estimates of the prevalence of DV during pregnancy in LMIC settings. The implementation of the tool was feasible and required participant convenience, particularly regular ANC visits rather than extensive resources. The positive reception of the study invitation among the women, despite the topic’s sensitive and taboo nature in the cultural context, promotes the acceptability of our approach. By prioritizing the perspectives of individual women, our research adopts a woman-centered approach that acknowledges the significance of their voices, particularly in societies wherein they may be marginalized or excluded. Nevertheless, the study has limitations that must be acknowledged so future research can aim to overcome them and expand the scope and inclusivity of studies on DV during pregnancy in Nepal.

First, the study was conducted in two largely urban hospitals, which may inaccurately reflect the prevalence of DV during pregnancy in all regions of Nepal; thus, further research is needed to encompass a wider range of healthcare settings and diverse geographic locations. Additionally, the small sample size precludes a focus on sub-group analysis, perpetrator type, with low prevalence. Second, linguistic barriers prevented several potential participants from participating in the study. Although Nepali is the official working language at the federal level, some individuals may have trouble understanding it, and this limitation may have led to the exclusion of individuals who could have provided valuable insights from their experiences of potential DV encounters. Thus, efforts should be made in future studies to address language barriers and ensure inclusivity. Finally, the study did not include participants with physical disabilities, such as poor vision and hearing, and the absence of sign language interpretation in the study materials may have hindered these individuals’ participation. As such, future research should consider accessibility measures to enable the inclusion of individuals with disabilities, as they may belong to vulnerable groups that could provide valuable perspectives on DV experiences.

## Conclusion

The N-AAS exhibited strong test-retest reliability, concurrent validity, specificity, positive predictive value, negative predictive value, likelihood ratio, and Kappa coefficient, where excellent sensitivity ratings were observed for physical abuse within the last 12 months and emotional and physical abuse since marriage produced moderate sensitivity ratings. Furthermore, sexual abuse, heavy work during pregnancy, physical abuse during pregnancy, and food deprivation warrant further investigation with a larger sample size, and participants should be assured that their responses will be published in a manner guaranteeing their anonymity.

## Data Availability

The datasets generated and analyzed in this study are not publicly available due to their sensitive nature given that release of the information could compromise the participants safety. An anonymized version of the data can be made available upon reasonable request. The data is secured with the first and second author in an encrypted pen drive in a locker.

## Acknowledgements

We are grateful to all participating pregnant women, the staff, and our research assistants at the antenatal clinics of Hospital 1 and Hospital 2, as well as to Prabin Raj Shakya for programming the C-ACASI.

The ADVANCE 2 research team includes the following people: *Norway–*Principal Investigator Professor Berit Schei, Norwegian University of Science and Technology (NTNU); Associate Professor Jennifer J. Infanti, NTNU; Professor Eva Skovlund, NTNU; Associate Professor Melanie Rae Simpson, NTNU; Co-principal Investigator Professor Mirjam Lukasse, University of South-Eastern Norway; Professor Lena Henriksen, Oslo Metropolitan University; Sweden*–*Professor Katarina Swahnberg, Linnæus University; USA*–*Professor Jacquelyn Campbell, John Hopkins University School of Nursing; Nepal*–*Principal Investigator Professor Sunil K. Joshi, Kathmandu Medical College (KMC); post-doctoral researcher Poonam Rishal, KMC; site principal investigators at Dhulikhel, Professors Rajendra Koju and Kunta Devi Pun; PhD candidates Pratibha Chalise and Pratibha Manandhar; and research assistants Kritika Shrestha, Usha Thapa, Nani Maiya Basi, Asma Pun, and Shanta Banjara.

## Ethics approval

The study was approved by the Regional Committee for Medical and Health Research Ethics Mid-Norway (178092) and by the ethics committees of the participating hospitals, as well as the Nepal Health Research Council (2395).

## Consent for publication

Not applicable.

## Availability of data and materials

All data and materials used in this study are available on request to corresponding author.

## Author contributions

Conceptualization, all authors; Data curation, PM, PC; Formal analysis, PM, PC; Funding acquisition, BS, JJI; Investigation, PM, PC; Methodology, PM, PC, JJI, BS, KS; Project administration, SKJ, KDP, JJI, KS; Supervision, PR, LH, JJI, SKJ, ML, KDP, KS; Writing— original draft, PM; Writing—review & editing, all authors. All authors have read and agreed to the final version of the manuscript.

